# ZIP code-level alcohol outlet density and nonfatal overdose among people who inject drugs in 22 US metropolitan statistical areas: a multilevel modeling analysis

**DOI:** 10.1101/2025.06.23.25330126

**Authors:** Snigdha R. Peddireddy, Stephanie Beane, Courtney Yarbrough, Umedjon Ibragimov, Janet R. Cummings, Danielle F. Haley, Sabriya L. Linton, Hannah L.F. Cooper

## Abstract

**Background:** Alcohol outlet density (AOD) is associated with drinking behaviors and related harms across several populations. As alcohol consumption compounds the depressive effects of opioids to increase overdose risk among people who inject drugs (PWID), this study investigated 1) whether off-premise AOD is associated with the risk of nonfatal overdose among a large sample of PWID, and 2) whether this relationship varies by individual race/ethnicity.

**Methods:** We linked individual-level 2018 National HIV Behavioral Surveillance (NHBS) data with ZIP code-level data on off-premise AOD in 2016 from the US Census Bureau’s ZIP Code Business Pattern survey. NHBS surveys PWID across 22 metropolitan statistical areas. Hierarchical generalized linear models quantified the association between AOD and nonfatal overdose, overall and by PWID race/ethnicity.

**Results:** The sample comprised 9,764 PWID who used opioids, 45% of whom were non-Hispanic/Latinx White, 38% non-Hispanic/Latinx Black, and 18% Hispanic/Latinx. Over a quarter (28%) experienced an overdose in the past year. The median outlet density was 4.0 outlets per square mile. The adjusted model did not show a significant relationship between AOD and odds of overdose (OR: 1.01, 95% CI: 1.00-1.02), and the relationship remained null in the race/ethnicity interaction model.

**Conclusions:** Regulating AOD alone may not effectively mitigate overdose risk among PWID. Future research should explore why overdoses among PWID are not sensitive to changes in AOD. Possible explanations to consider are whether (1) AOD is unassociated with alcohol consumption among PWID, or (2) PWID stagger the timing of their alcohol consumption so it does not coincide with opioid consumption.

## INTRODUCTION

Provisional drug overdose data from the United States (US) indicate that there were over 81,000 opioid-involved overdose deaths in 2023 [1]. While most recent research has focused on stimulant co-involvement in the latest wave of the opioid epidemic, alcohol has also played a significant role in opioid overdoses: alcohol was involved in approximately 17% of opioid overdose deaths in the US in 2020, up 41% from 2019 [2]. The concurrent consumption of alcohol and opioids has synergistic effects that amplify the respiratory depressant properties of both substances, and significantly elevate the risk of overdose [3,4].

Evidence suggests that alcohol consumption in the general population is influenced, in part, by the density of off-premise alcohol outlets (i.e., outlets where alcohol is sold for consumption off retail outlet property, such as grocery and convenience stores). Off-premise alcohol outlet density (AOD) has consistently been linked to harmful drinking behaviors and related acute harms [5–8]. This body of evidence has supported the regulation of AOD as a means to mitigate these risks in the general population [8–10]. AOD regulation typically involves zoning and licensing policies, such as limiting areas where alcohol sales are allowed or setting limits on the number of licensed outlets according to set criteria, respectively, enabling local authorities to reshape neighborhood alcohol risk environments [11].

Despite the established connection between AOD and alcohol consumption in the general population, and between alcohol consumption and overdoses among people who use drugs, exploration into AOD’s relationship with drug overdose has been limited. To date, only one study has investigated this relationship: an ecologic study found that among census block groups within Baltimore City, each additional off-premise outlet was associated with a 16.6% increase in the rate of overdoses from any drugs among the general population of residents [12]. People who inject drugs (PWID) face a heightened risk of overdose compared to people who consume opioids through other administration routes, as injecting introduces drugs into the bloodstream more immediately and with greater potency [13]. While Nesoff et al. (2021) explored the relationship between AOD and overdose among the general population at the neighborhood level [12], it remains uncertain how these findings extend specifically to PWID.

Additionally, ecological studies and aggregate data may obscure differential impacts on marginalized subgroups. Specifically, the same level of exposure to alcohol outlets may have more deleterious impacts on racially minoritized individuals. Between 2017 and 2020 in Illinois, the prevalence of alcohol co-involvement in opioid overdose deaths was 39% and 33% among Hispanic/Latinx and Black decedents, respectively, compared to 27% among White decedents [14]. The increased availability of alcohol in racialized alcohol risk environments may intensify already growing opioid overdose rates among racially minoritized individuals [15–17]. This study thus aimed to explore whether 1) off-premise AOD is associated with the risk of nonfatal overdose among a large sample of PWID and 2) the relationship between AOD and overdose varied by individual race/ethnicity.

## METHODS

### Overview

This analysis utilized data from the Center for Disease Control and Prevention’s (CDC) National HIV Behavioral Surveillance (NHBS) study, the largest and most racially/ethnically diverse source of health data from PWID across multiple metropolitan statistical areas (MSAs) in the US. The CDC implemented NHBS to monitor behavioral risk and protective factors associated with HIV. In 2018, NHBS aimed to gather data from approximately 500 PWID in each of the 23 MSAs using respondent-driven sampling (RDS).

This multilevel cross-sectional study linked 2018 NHBS data on 9,764 individual PWID with ZIP code-level AOD data from the US Census Bureau’s 2016 ZIP Code Business Pattern (CBP) survey. Complete AOD data after 2016 was unavailable due to CBP’s suppression of cells for ZIP codes with fewer than three establishments after this year. We thus linked the most recent and complete AOD data in 2016 with 2018 NHBS data (two-year lag) to investigate potential temporal relationships.

### Sample

PWID were recruited to the NHBS study if they (1) reported injecting a non-prescribed drug in the past 12 months, (2) were aged 18 years or older, (3) lived in a participating MSA, (4) had not previously participated in the current cycle of data collection, and (5) could complete the survey in English or Spanish. This analysis further restricted the sample to participants who reported past-year injection or non-injection use of a non-prescribed opioid (99% of the initial sample). The analytic sample excluded 1) participants who reported a race/ethnicity other than Black, non-Hispanic/Latinx; Hispanic/Latinx; or White, non-Hispanic/Latinx due to sparse data for other races/ethnicities and 2) participants with missing values for variables used in this analysis, including missing ZIP codes (< 5% of the sample). Participants from the San Juan-Bayamon MSA were excluded as well because of a lack of racial/ethnic diversity, resulting in 22 MSAs represented in the analytic sample.

### Measures

#### Outcome

##### Past 12-month non-fatal opioid overdose

The outcome was a dichotomous variable indicating whether the NHBS participant experienced a non-fatal opioid overdose in the past 12 months based on the following question: “In the past 12 months, did you overdose on heroin or painkillers? By overdose, I mean if you passed out, turned blue, or stopped breathing from using drugs.”

#### Primary independent variable

##### ZIP code-level alcohol outlet density

The primary independent variable was the ZIP code-level density of off-premise alcohol outlets, calculated as the number of outlets divided by the square miles in each Zip Code Tabulation Area (ZCTA), defined as the US Postal Service’s ZIP code service areas [18]. Data on the numerator were obtained from the 2016 U.S. Census Bureau’s ZIP Code Business Pattern (CBP) data [19]; the two-year lag allowed for delayed impacts on the outcome. The CBP reports annual, sub-national economic data on business establishments by industry type. ZIP-code level CBP data included the number and type of establishments (supermarkets/grocery stores, convenience stores, beer/wine/liquor stores, gas stations with convenience stores, and pharmacies/drug stores) that could potentially sell alcohol for off-premise consumption according to each state’s liquor licensing laws. PWID were linked to 2016 AOD using the reported ZIP code of residence.

#### Covariates

##### State-level covariates

*Alcohol policy scores* quantified the strengths of states’ alcohol policy landscapes in 2017 [20]. Scores ranged from 0-100 and higher scores represented more comprehensive and restrictive alcohol environments. Indicators included policies regarding alcohol production and distribution, sales, and consumption. PWID were linked to states using the reported county of residence.

##### ZIP code-level covariates

We used 2017 (one-year lag) United States Postal Services Address Information Systems data [21] to construct *1) residential and 2) business vacancies* by dividing the monthly average of vacancies in each category by ZCTA square miles. Residential and business vacancies shape excessive drinking and drug-related behaviors, as well as opioid-related overdoses [22,23]. We also calculated 2017 (one-year lag) *percent of people with income at or below the federal poverty line* using the midpoint of the U.S. Census Bureau’s American Community Survey 5-year block data [24]. We did not include a measure of racial segregation in our models due to potential multicollinearity with AOD, as indicated by the racialized concentration of alcohol outlets [25–30].

##### Individual-level covariates

Variables capturing individual-level characteristics were drawn from NHBS. These included participant sociodemographic characteristics (race/ethnicity, sex, age, education, employment), poverty status per the US DHHS Poverty Guidelines [31], health insurance status, daily injection drug use, incarceration (past 12 months), unhoused status (past 12 months), HIV serostatus, self-reported disability status using the DHHS data standard items [32,33], psychological distress as measured by the Kessler-6 scale [34], network size, respondent status as an RDS seed, and receipt of sterile injection drug use equipment from a syringe services program in the past year to reflect access to harm reduction services and treatment, associated with our overdose outcome [35,36].

#### Potential effect modifier

##### Individual race/ethnicity

NHBS participants’ self-reported data informed three mutually exclusive racial/ethnic groups: Hispanic/Latinx; White (hereinafter ‘White’); and non-Hispanic/Latinx Black (hereinafter ‘Black’). When participants reported that they belonged to two racial groups and were not Hispanic/Latinx, this study followed the Office of Management and Budget’s “plurality” guidelines to assign them to a single racial/ethnic group using the federal Office of Management and Budget’s “plurality” guidelines [37].

### Analysis

Statistical analyses were carried out with SAS version 9.4. We first explored distributions of all variables, examined correlations, and checked for multicollinearity. Age and number of years since first injection were highly correlated (0.75), so injection duration was removed from all models. We conducted a cross-tabulation and chi-square test to discern statistically significant differences in past 30-day binge drinking among individuals who either experienced or did not experience an overdose in the past year. We also compared median alcohol outlet densities by individual PWID race/ethnicity using a Kruskal-Wallis test [38].

Three-level hierarchical generalized linear models with a logit link [39] examined the associations between ZIP code-level AOD and nonfatal opioid overdose. Models included random intercepts for PWID clustered within ZIP codes and ZIP codes clustered within MSAs. Models could not support a random intercept for both MSA and state, as only three states contained more than one MSA (unconditional models with MSAs and states resulted in a state-level variance component of zero [p > .05] and a warning that the estimated G matrix was not positive definite). Previous work has shown that when the highest level (here, state) of clustering is ignored in a model, that level’s variance component is redistributed to the lower level [40], so models only included a random intercept for MSA as the highest spatial level of clustering.

Three models are reported: 1) an unadjusted model, 2) a multivariable model adjusted for state, ZIP code, and individual PWID covariates and 3) an adjusted multivariable model with interactions to explore whether the relationship of AOD to nonfatal opioid overdose varied by race/ethnicity. We express model outcomes as odds ratios for overdose where the AOD is set to increments of three outlets per square mile above the median AOD (equivalent to about half of the interquartile range). This approach facilitates comprehension of the relationship between a higher AOD and the odds of nonfatal overdose. Additionally, we based AOD increments on the median, as the distribution of AODs was highly skewed in our sample. While we report the prevalence of binge drinking in our sample for descriptive purposes, this measure was not included in the models as it is a potential mediator in the relationship between AOD and overdose.

## RESULTS

### Sample description

The sample of 9,764 PWID lived in 20 states and 22 MSAs. Approximately two-thirds (68%) of the sample were male; 45% were White, 38% were Black, and 18% were Hispanic/Latinx (Table 1). The majority of participants exhibited indicators of socioeconomic hardship: approximately 75% subsisted at or below the federal poverty guidelines, 59% were unemployed, and 25% were unable to work due to poor health. Regarding health indicators, 89% reported daily injection drug use, 39% experienced psychological distress in the past month, and 68% had a qualifying disability. Importantly, 28% of participants reported experiencing an opioid overdose in the past year. The prevalence of binge drinking among our sample (27%) was similarly high. Among PWID who had overdosed in the past year, 31% reported binge drinking in the past month, and, among those who did not overdose, 26% reported binge drinking. This difference was statistically significant (χ^2^= 26.4, p < 0.0001).

**Table 1:**
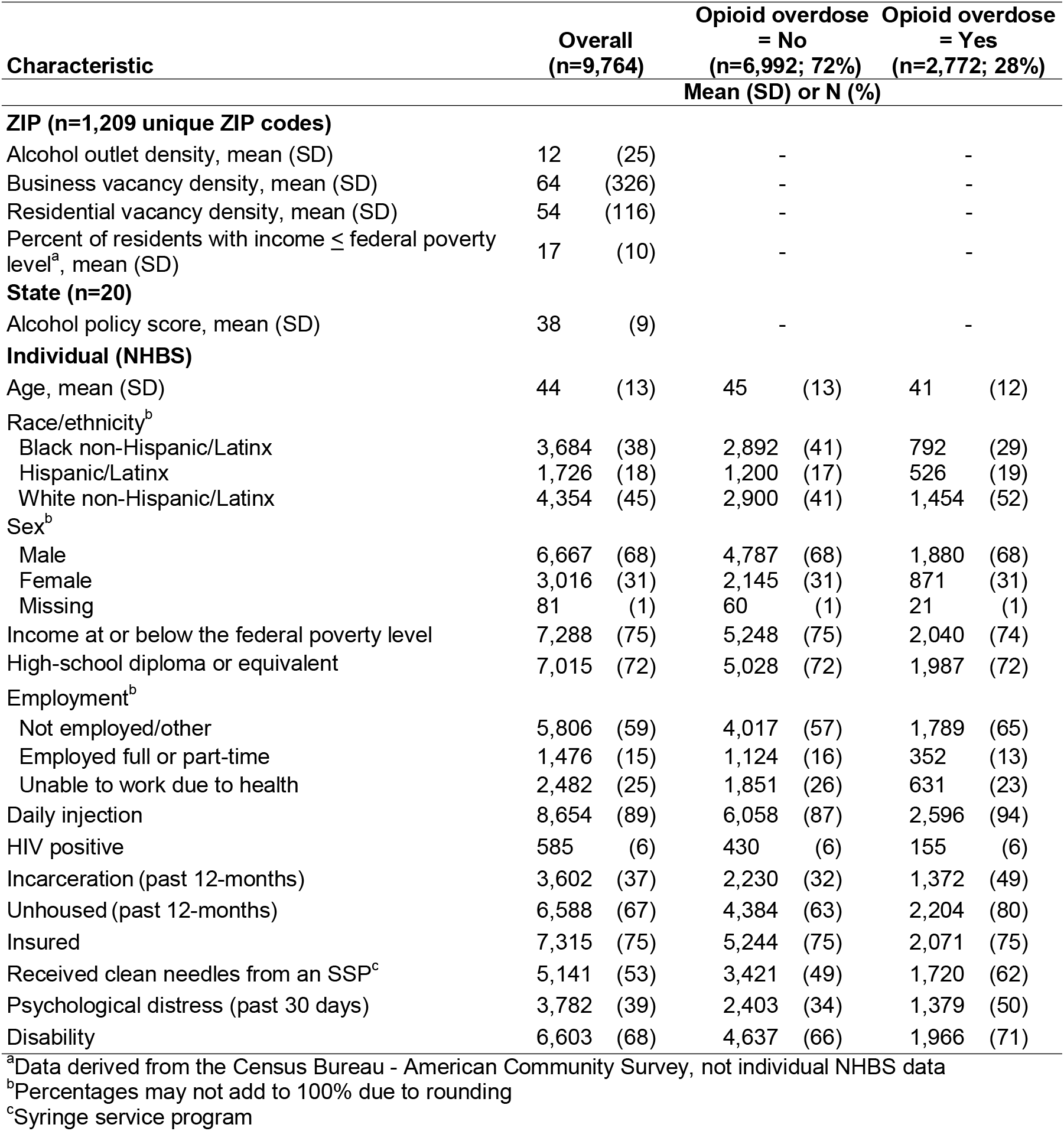
ZIP, state, and participant characteristics, overall and by opioid overdose, 2018 Centers Disease Control and Prevention’s National HIV Behavioral Surveillance (NHBS) data (n=9,764)

Among the 1,209 unique ZIPs where NHBS PWID resided, the mean AOD was 12 (SD=25) off-premise alcohol outlets per square mile, and the median was 4.0 (range: 0-266.16); the distribution of AODs among our sample was positively skewed.

### Model-based analysis

In the unadjusted model, as ZIP code-level AOD increased by three outlets per square mile above the median AOD (4.0), the odds of overdose among PWID in our sample increased by 2% (OR: 1.02; 95% CI: 1.01-1.03, p = 0.003) (Table 2). However, the magnitude of this relationship was attenuated to non-significance (OR: 1.01, 95% CI: 1.00-1.02, p = 0.23) in the adjusted model controlling for state-level alcohol policy scores and ZIP code- and individual-level covariates. In the race/ethnicity interaction model, interaction terms were not statistically significant (p = 0.16 – 0.17).

**Table 2:** Unadjusted and adjusted hierarchical generalized linear regressions^a^ of the odds of non-fatal opioid overdose on alcohol outlet density and race/ethnicity potential effect modifier, 2018 Centers Disease Control and Prevention’s National HIV Behavioral Surveillance (N=9,764)

|  | OR (95% CI), p value |
| --- | --- |
| **Unadjusted model** |  |
| Alcohol outlet density<sup>b</sup> | 1.02 (1.01, 1.03), 0.003 |
| Var (SE) for MSA | 0.15 (0.05), 0.002 |
| Var (SE) for ZIP | 0.05 (0.02), 0.006 |
| **Adjusted<sup>c</sup> model** |  |
| Alcohol outlet density<sup>b</sup> | 1.01 (1.00, 1.02), 0.23 |
| Race/ethnicity (Ref: White non-Hispanic/Latinx) |  |
| Black non-Hispanic/Latinx | 0.80 (0.70, 0.91), < 0.001 |
| Hispanic/Latinx | 0.96 (0.83, 1.10), 0.53 |
| Var (SE), p value for MSA | 0.11 (0.04), 0.004 |
| Var (SE), p value for ZIP | 0.007 (0.01), 0.30 |
| **Adjusted model<sup>c</sup> including interaction with race/ethnicity** |  |
| Alcohol outlet density<sup>b</sup> | 1.01 (1.00, 1.03), 0.06 |
| Race/ethnicity (Ref: White non-Hispanic/Latinx) |  |
| Black non-Hispanic/Latinx | 0.83 (0.71, 0.95), 0.009 |
| Hispanic/Latinx | 1.00 (0.85, 1.16), 0.97 |
| Alcohol outlet density * race/ethnicity, ORR<sup>d</sup> |  |
| Black non-Hispanic/Latinx | 0.99 (0.99, 1.002), 0.17 |
| Hispanic/Latinx | 1.00 (0.99, 1.001), 0.16 |
| Var (SE), p value for MSA | 0.10 (0.04), 0.004 |
| Var (SE), p value for ZIP | 0.007 (0.01), 0.30 |

<sup>a</sup>3-level (Level 1: Individual PWID, Level 2: ZIP, Level 3: MSA) hierarchical generalized linear model with logit link
<sup>b</sup>OR offset 3 (approximately half the interquartile range) from the median alcohol outlet density for unique ZIP codes (4.04)
<sup>c</sup>Model adjusted for state-level alcohol policy score, ZIP-level covariates (density of business and residential vacancies, percent of residents with income at or below the federal poverty level), and individual covariates (race/ethnicity, sex, age, income, education, employment, health insurance, incarceration, daily injection status, unhoused status, HIV status, network size, receipt of clean needles from an SSP, psychological distress, and disability status)
<sup>d</sup>Odds ratio ratio

As a sensitivity analysis, we ran models excluding alcohol policy scores. The interpretation of the relationship between AOD and overdose was unchanged in sensitivity analysis models (no or negligible change to estimates and confidence intervals).

## DISCUSSION

Leveraging the CDC’s comprehensive NHBS data allowed this analysis to generate three important findings. First, a high prevalence of nonfatal opioid overdose was observed among PWID—with more than one in four participants reporting an overdose in the past year— and a higher percentage of those who had overdosed reported binge drinking compared to those who had not overdosed. Second, while AOD and overdose were significantly associated in the unadjusted model, this relationship did not persist after adjusting for potential confounders. Lastly, the relationship between AOD and overdose did not vary by race/ethnicity.

In contrast to Nesoff et al’s (2021) finding that off-premise AOD was positively associated with neighborhood-level nonfatal overdose rates among Baltimore residents [12], we did not identify a significant relationship between outlet density and self-reported nonfatal overdose among *PWID* after adjusting for multilevel factors. Several reasons may explain these disparate findings. We were able to analyze this relationship among PWID specifically, rather than the general population, and PWID may carefully manage the timing of their alcohol consumption in relationship to their opioid consumption. Insights from a qualitative study of PWID lend credence to this possibility: Edsall et al. found that PWID avoid alcohol while using heroin to 1) mitigate riskier heroin use that could follow alcohol consumption and thus lead to an increased risk of overdose, and 2) to avoid alcohol’s blunting of opioid’s effects [41]. PWID may be more knowledgeable about overdose prevention—including about alcohol’s amplification of opioid overdose rise—than people who primarily use drugs through other administration routes [42,43]. We could not examine these potential confounders.

Another potential explanation for the lack of a significant relationship between AOD and nonfatal overdose in our study, as opposed to Nesoff et al.’s city-specific findings [12], might be the nationwide scope of our analysis. Investigating 22 MSAs as opposed to a single city could introduce heterogeneity that obscures any distinct, localized associations. Additionally, our study necessarily focused on nonfatal overdoses, because overdose was ascertained via self-report, while Nesoff analyzed EMS call counts on fatal and nonfatal overdose [12]. Possibly, AOD is associated with higher rates of fatal overdose because of alcohol’s amplifying effects on respiratory suppression.

We must consider several limitations of our analysis. The utilization of 2018 NHBS data, combined with 2016 AOD data, does not allow us to consider more recent trends in overdose and alcohol availability. Recent data have indicated that overdoses among Black Americans now outpace those among their White counterparts [15–17]. Also, there has been a substantial increase in alcohol availability during the COVID-19 pandemic due to the expansion of home delivery services, requiring that we expand the definition of off-premise availability [44]. As a result, our estimates of the rates of overdose and of outlet density, a measure of alcohol availability that only accounts for brick-and-mortar establishments and not online or third-party delivery services, may be conservative. Recent studies have found a greater magnitude of the effect of off-premise alcohol availability on alcohol consumption and related harms after the onset of the COVID-19 pandemic [45,46]; thus, our estimate for the association between off-premise outlet density and overdose may also be conservative. Similarly, CBP data suppress outlets with fewer than ten employees, leading to underreporting of ZIP code-level AOD. The NHBS survey did not ascertain alcohol co-involvement in nonfatal overdoses or alcohol and opioid co-use patterns, so we were not able to account for these factors in our analysis. Self-reported overdose data may have also led to an underestimate of our outcome. Another limitation is the use of ZIP codes as the primary spatial unit of analysis. ZIP codes may not accurately reflect substance use behaviors within individual census tracts or other more granular levels of analysis, where drug-related harms occur. Also, 67% of participants in this study’s sample were unhoused in the previous year, and reported residential ZIP codes may not have aligned with their actual activity spaces (i.e., the areas where they most frequently accessed or consumed alcohol). Lastly, findings are not generalizable to all PWID in the US, as participants were recruited from only major with high HIV/AIDS prevalences; likewise, findings may not be generalizable to the underlying population of PWID in the studied MSAs.

In sum, while some evidence points toward the regulation of AOD as a potential avenue to reduce overdose risk in the general population, this regulation alone may not address overdose risk among PWID specifically. However, while AOD may not be directly associated with overdose risk among PWID, it may still be related to other adverse outcomes among PWID that warrant further investigation (e.g., binge drinking, mental health distress, risky sexual behaviors). Future research should delve deeper into why AOD may not be linked to overdose among PWID as it is in the general population, exploring factors such as PWID’s intentional timing of alcohol consumption relative to injection drug use, and whether specific patterns of co-use (e.g., binge drinking while injecting) might influence overdose risk.

## Data Availability

All data produced and analyzed in the present study are only available upon request to the Centers for Disease Control and Prevention's National HIV Behavioral Surveillance program.

## ABBREVIATIONS

AOD: alcohol outlet density
CBP: ZIP Code Business Pattern
MSA: metropolitan statistical area
NHBS: National HIV Behavioral Surveillance
ZCTA: ZIP code tabulation area

## DECLARATIONS

## Ethics approval and consent to participate

This study was approved by the Emory University Institutional Review Board.

## Consent for publication

Not applicable

## Availability of data and materials

The data used in this study are not publicly available due to Centers for Disease Control and Prevention (CDC) data use restrictions. The study team obtained access to the data through a guest researcher partnership with the CDC, which precludes us from sharing the data directly. Any data requests must be made directly to the CDC.

## Authors’ contributions

**SP:** Conceptualization, Methodology, Writing – Original Draft, Writing – Review & Editing, **SB:** Methodology, Software, Formal Analysis, Data curation, Writing – Original Draft, Writing – Review & Editing, **CY, UI, JC, DH, SL:** Writing – Reviewing & Editing, **HC:** Supervision, Funding Acquisition, Writing – Reviewing & Editing. All authors reviewed the final manuscript.

## Acknowledgements

We express deep appreciation to the reviewers and Editor at *Harm Reduction Journal* and our funders. We are also grateful to Jason Blanchette, JD, MPH (Boston University School of Public Health) and Tim Naimi, MD, MPH (University of Victoria) for providing the alcohol policy scores used in this analysis. Lastly, we would like to express our gratitude to the NHBS PWID participants and to the Principal Investigators and teams at the NHBS sites, including: Atlanta, GA: Pascale Wortley, Jeff Todd, David Melton; Baltimore, MD: Colin Flynn, Danielle German; Boston, MA: Monina Klevens, Rose Doherty, Conall O’Cleirigh; Chicago, IL: Antonio D. Jimenez, Thomas Clyde; Dallas, TX: Jonathon Poe, Margaret Vaaler, Jie Deng; Denver, CO: Alia Al-Tayyib, Daniel Shodell; Detroit, MI: Emily Higgins, Vivian Griffin, Corrine Sanger; Houston, TX: Salma Khuwaja, Zaida Lopez, Paige Padgett; Los Angeles, CA: Ekow Kwa Sey, Yingbo Ma, Hugo Santacruz; Memphis, TN: Meredith Brantley, Christopher Mathews, Jack Marr; Miami, FL: Emma Spencer, Willie Nixon, David Forrest; Nassau-Suffolk, NY: Bridget Anderson, Ashley Tate, Meaghan Abrego; New Orleans, LA: William T. Robinson, Narquis Barak, Jeremy M. Beckford; New York City, NY: Sarah Braunstein, Alexis Rivera, Sidney Carrillo Newark, NJ: Abdel R. Ibrahim, Afework Wogayehu, Luis Moraga; Philadelphia, PA: Kathleen A. Brady, Jennifer Shinefeld, Chrysanthus Nnumolu,; Portland, OR: Timothy W. Menza, E. Roberto Orellana, Amisha Bhattari; San Diego, CA: Anna Flynn, Onika Chambers, Marisa Ramos; San Francisco, CA: Willi McFarland, Jessica Lin, Desmond Miller; San Juan, PR: Sandra Miranda De León, Yadira Rolón-Colón, María Pabón Martínez; Seattle, WA: Tom Jaenicke, Sara Glick; Virginia Beach, VA: Jennifer Kienzle, Brandie Smith, Toyah Reid; Washington, DC: Jenevieve Opoku, Irene Kuo.

